# Determinants of Low Birth Weight Babies Delivered at Paropakar Maternity and Women’s Hospital: A Case-Control Study

**DOI:** 10.1101/2025.10.08.25337614

**Authors:** Sadiksha Pokhrel, Paras Kumar Pokharel, Anup Ghimire, Birendra Kumar Yadav, Avaniendra Chakravartty, Swastika Poudel, Laxmiswori Prajapati, Akshaya Acharya

**Author notes:** **Corresponding Author:** (SP).

## Abstract

**Background:** Low birth weight (LBW), defined as infants weighing less than 2500 grams, is a key indicator of health and developmental outcomes. It is associated with long term consequences such as increased risks of behavioral, psychological and learning challenges. The contributing factors of LBW includes poverty, maternal smoking, limited access to healthcare and so on. Understanding these influences is essential for effective intervention strategies. Thus, this study aimed to identify the risk factors associated with low birth weight babies delivered at Paropakar Maternity and Women’s Hospital in Kathmandu, Nepal.

**Methods:** Hospital based age matched (± 5 years) case-control study was carried out from 1^st^ August 2024 to 30^th^ January 2025 among the mothers who delivered live babies at Paropakar Maternity and Women’s Hospital. A total of 57 cases and 114 controls in the ratio of 1:2 were selected as study participants. Information regarding exposure status was obtained through interviews and medical records. Chi-square test, mann-whitney U test and binary logistic regression were performed at 95% CI to test the association by using SPSS version 11.5.

**Results:** The adjusted multivariate logistics regression analysis revealed that independent risk factors associated with LBW baby were educational status of mother (AOR: 6.32; 95% CI 1.90 to 21.05), per capita income (AOR: 2.89; 95% CI 1.02 to 8.18), parity (AOR: 4.80; 95% CI 1.91 to 12.07), hemoglobin level (AOR: 6.19; 95% CI 1.79 to 21.38), period of gestation (AOR: 8.16; 95% CI 2.42 to 27.49), weight before pregnancy (AOR: 4.86; 95% CI 1.02 to 23.29), history of chronic medical illness (AOR: 8.22; 95% CI 2.25 to 29.99) and illness during pregnancy (AOR: 3.33; 95% CI 1.20 to 9.28), type of diet (AOR: 4.84; 95% CI 1.14 to 20.64)

**Conclusion:** Study findings suggest that selected targeted interventions such as improving maternal education and nutrition, intake of iron rich food, increasing maternal weight before pregnancy, avoiding smoking and alcohol consumption during pregnancy can prevent LBW in Nepal.

## Introduction

WHO had defined low birth weight as the weight of infants less than 2500 grams at birth irrespective of the gestational age.^(1)^ The proportion of babies with LBW is considered as a sensitive index that represents the country’s health and development. ^(2)^ The babies with LBW are at a greater risk of malnutrition as well as other childhood morbidities such as pneumonia and diarrhea that are considered as the major challenging public health problem. ^(3)^ LBW infants lack the strength as compared to the babies born with normal weight due to their tiny bodies and are at more risk to various health related problems. As they lack body fat, they typically struggle to stay warm and might suffer from hypothermia. The baby’s risk for complications increases with the low birth weight.^(4–6)^

LBW babies are more likely to die during the first month of their life. They might face lifelong consequences including a higher risk of stunted growth, lower IQ, and adult-onset chronic conditions such as hypertension, ischemic heart disease, stroke, metabolic syndrome, obesity, diabetes, malignancies, osteoarthritis and dementia.^(7,8)^ It is also associated to an increased risk of behavioral problems, psychological illnesses, learning difficulties, and sensory impairments that affect cognitive function in developing children and adolescents that provide significant challenges to the individual’s educational and quality of life outcomes.^(2)^

It has been estimated that 15-20% of infants worldwide weigh less than 2,500 grams at birth, which is equal to more than 20 million births annually.^(9)^ Worldwide, one out of seven newborns are affected due to LBW.^(7)^ Out of total LBW babies worldwide, 95% occur in developing nations, which is more than twice as high as in developed nations i.e. 16.5% and 7.0% respectively.^(2,10)^ Southern Asia (27%) has the highest incidence of LBW followed by Africa (14%), Latin America and the Caribbean (9%), and Eastern Asia (6%). ^(8,11,12)^ Based on the data from Global Nutrition Report, 12.3% of newborns in the South-eastern Asia sub-region have low birth weight.^(13)^

In Nepal, the percentage of newborns with low birth weight has slightly decreased from 12.8% in FY 2076/77 to 11.2% in this FY 2077/78. According to Nepal Demographic Health Survey (NDHS) 2016, the prevalence of low birth weight is 12%. Likewise, the data from the NDHS 2022 reported that 5% of babies were very small at birth and 10% were smaller than average. Based on these data, during the past decade, the proportion of newborns with low birth weight has remained stagnant.^(14,15)^ LBW is influenced by a complex interplay of socio-demographic and environmental factors, including poverty, maternal smoking, and inadequate access to healthcare. Understanding the role of these factors in the context of Nepal is essential for designing effective interventions. This research has the potential to improve maternal and child health outcomes, reduce neonatal mortality, and contribute to achieving Sustainable Development Goal 3 while addressing the unique challenges and regional variations within Nepal by identifying the modifiable risk factors that are associated with the delivery of low birth weight baby.

Despite several studies on low birth weight (LBW) in Nepal, evidence from Kathmandu Valley is limited. Paropakar Maternity and Women’s Hospital, the country’s largest referral center, serves mothers from both urban and rural backgrounds, offering a unique setting to study diverse risk factors. With LBW rates in Nepal showing little improvement in the past decade (NDHS 2022), updated hospital-based evidence from this context is essential to guide effective interventions. Thus, this study aimed to identify the risk factors associated with low birth weight babies delivered at Paropakar Maternity and Women’s Hospital in Kathmandu, Nepal.

## Materials and Methods

### Methods

Hospital-based age matched case-control design (age of mothers +/− 5 years) was employed in this study to identify the determinants of low birth weight baby. Paropakar Maternity and Women’s Hospital, Kathmandu, was purposively selected as a study setting. Data collection was done from 1^st^ August 2024 to 30^th^ January 2025.

### Participants

#### Cases

Those women delivering the newborn with birthweight less than 2500 gram at Paropakar Maternity and Women’s Hospital.

#### Controls

Those women delivering the newborn with birthweight greater than or equal to 2500 gram at Paropakar Maternity and Women’s Hospital.

### Inclusion and exclusion criteria

All those women who delivered singleton live baby at the hospital during the data collection period were included in the study while those women who gave birth to twin babies were excluded.

### Sample size and sampling technique

The number of cases and controls were calculated by using OpenEpi, Version 3 by taking; power at 80%, two sided significant level at 0.05, percentage of controls exposed 35.41 and odds ratio of 2.51 by taking gestational age as a exposure variable based on the study conducted by Shrestha et.al (2020) at Lumbini Provincial Hospital.^(16)^ The ratio of case to control is taken at 1:2. Thus, the required sample size calculated was 57 cases and 114 control.

Delivery record book was checked for the delivery of low birth weight baby each day during the data collection period in the Post Natal Care ward of Paropakar Maternity and Women’s Hospital. Mother who gave birth to LBW baby (birth weight <2500gm) were taken as cases for the data collection. Similarly, two controls (birth weight ≥2500gm) were selected on the same day when a case was found for the data collection by matching age with case.

### Validity and reliability

To ensure the validity and reliability, the questionnaire to assess the risk factors of LBW babies was prepared from extensive literature review. The prepared structured questionnaire was translated into the Nepali language and then back-translated into English language to ensure validity and reliability of the questionnaire. Pre-testing of the questionnaire was done for 10% of study samples in a similar setting which were not included in the main study and required changes were made accordingly.

### Data collection tools and techniques

The pre-tested structured standard questionnaire was used to conduct the study. Face-to-face interview using the structured questionnaire was conducted for data collection. Some medical and obstetric information obtained from the participants were cross verified by reviewing the medical records of the participants.

### Variables

Independent variables: Socio-demographic characteristics, Maternal and obstetric factors, and Nutritional and behavioral factors of mothers.

Outcome variable: Low birth weight baby as defined by birth weight < 2500 gram.

### Statistical analysis

Data was collected in paper-based questionnaire and entered in Microsoft excel 2010 which was exported to statistical package for social sciences (SPSS) 11.5 version for statistical analysis. Descriptive and inferential statistical methods were used to analyze and compare various characteristics among mothers of case and control group, and to identify associations between various factors and delivery of low birth weight baby. The Kolmogorov – Smirnov test and Shapiro – Wilk test, Q-Q plot, Skewness and Kurtosis, Histogram and Box and Whisker plot were conducted to evaluate whether the data was normally distributed or not.

For the descriptive analysis, categorical data of both cases and controls were summarized in terms of frequency and percentage. The continuous variables were presented as Median and Inter-Quartile Range (IQR).

For the inferential statistics, bivariate analysis was done using chi-square test and mann-whitney U test to determine factors associated with low birth weight baby at 95% CI and p-value less than 0.05. Those variables with p-value less than 0.2 in bivariate analysis were subjected for multivariate analysis. Binary logistic regression was used to find out significant factors associated with low birth weight after adjusting all possible confounding factors.

### Operational definitions

#### Per capita income

It has been categorized as below poverty line and above poverty line based on Nepal Living Standards Survey IV 2022-23. It was estimated at NRs.72,908 per person per year.(17)

#### MUAC

It is categorized as <23 cm and ≥ 23 cm based on Food and nutrition technical assistance (FANTA).(18)

#### Dietary diversity

It is categorized as adequate diverse diet and inadequate diverse diet. Women consuming 5 of 10 food groups in the past 24 hours while they were pregnant are defined as consuming a diverse diet using the Minimum Dietary Diversity Score for Women (MDD-W) tool given by FAO (Food and Agriculture Organization).^(19)^

#### Chronic medical illness

Chronic medical illness is defined as a pre-existing medical illness of the mother an onset prior to the current pregnancy.

#### Illness during pregnancy

Illness during current pregnancy is defined as a medical condition that developed during current pregnancy for which medical attention or treatment was sought.

## Results

### Socio-demographic characteristics of respondents

Table 1 represents the socio-demographic characteristics of the respondents. More than half of the newborns were male (53.2%). About one-third of the respondents were janajati (33.3%), more than two-third of the respondents followed Hindu religion (69.6%) followed by Buddhist religion (15.2%). About 13% of the mothers were illiterate and most of the mothers were home maker (69.6%). More than half of the respondents belonged to nuclear family (51.5%) and 76.6% of the respondents lived in a family with less than five members. The median monthly family income of the respondents was NRs 36,000 and about one-fourth of the participants were below poverty line (23.98%).

**Table 1.** Socio demographic characteristics of respondents.

| **Variables** | **Case (57)**<br>**n (%)** | **Control (114)**<br>**n (%)** | **Total (171)**<br>**n (%)** |
| --- | --- | --- | --- |
| **Sex of child** |  |  |  |
| Female | 30 (53.6%) | 50 (43.9%) | 80 (46.8%) |
| Male | 27 (47.4%) | 64 (56.1%) | 91 (53.2%) |
| **Ethnicity** |  |  |  |
| Brahmin | 17 (29.8%) | 17 (14.9%) | 34 (19.9%) |
| Chhetri | 9 (15.8%) | 19 (16.7%) | 28 (16.4%) |
| Newar | 7 (12.3%) | 10 (8.8%) | 17 (9.9%) |
| Dalit | 6 (10.5%) | 16 (14.0%) | 22 (12.9%) |
| Janajati | 15 (26.3%) | 42 (36.8%) | 57 (33.3%) |
| Others (Muslim, Madhesi) | 3 (5.3%) | 10 (8.8%) | 13 (7.6%) |
| **Religion** |  |  |  |
| Hindu | 41 (71.9%) | 78 (68.4%) | 119 (69.6%) |
| Buddhist | 10 (17.5%) | 16 (14%) | 26 (15.2%) |
| Muslim | 2 (3.5%) | 5 (4.4%) | 7 (4.1%) |
| Kirat | 1 (1.8%) | 8 (7%) | 9 (5.3%) |
| Christian | 3 (5.3%) | 7 (6.1%) | 10 (5.8%) |
| **Educational status of mother** |  |  |  |
| Illiterate | 12 (21.1%) | 10 (8.8%) | 22 (12.9%) |
| Literate | 45 (78.9%) | 104 (91.2%) | 149 (87.1%) |
| **Occupation of mother** |  |  |  |
| Agriculture | 2 (3.5%) | 6 (5.3%) | 8 (4.7%) |
| Government Job | 1 (1.8%) | 3 (2.6%) | 4 (2.3%) |
| Business | 5 (8.8%) | 8 (7.0%) | 13 (7.6%) |
| Home maker | 40 (70.2%) | 79 (69.3%) | 119 (69.6%) |
| Private Job | 7 (12.3%) | 13 (11.4%) | 20 (11.7%) |
| Daily wage | 2 (3.5%) | 5 (4.4%) | 7 (4.1%) |
| **Family size** |  |  |  |
| ≥ 5 members | 16 (28.1%) | 24 (21.1%) | 40 (23.4%) |
| < 5 members | 41 (71.9%) | 90 (78.9%) | 131 (76.6%) |
| **Monthly income (in NRs)** |  |  |  |
| Median (IQR) | 35000<br>(25000 - 50000) | 40000<br>(28750 - 50000) | 36000<br>(25000 - 50000) |
| **Per capita income** |  |  |  |
| Below Poverty Line | 20 (35.1%) | 21 (18.4%) | 41 (23.98%) |
| Above Poverty Line | 37 (64.9%) | 93 (81.6%) | 130 (76.02%) |

### Maternal and obstetric factors of respondents

Out of 171 respondents, more than half of the respondents had normal delivery (56.1%) and 52.6% of the respondents were primiparous. Majority of the respondents had inter-pregnancy interval of more than or equal to two years (88%) and had ANC visits for more than or equal to four times. About 13 % of the respondents had hemoglobin level ≥ 11 mg/dl. The period of gestation was less than 37 weeks among 14.6% of the respondents. More than one-fourth of the respondents had previously given birth to low birth weight babies (26.3%), 11.7% had abortion and 8.8% had history of preterm birth. About 11% of the respondents had height less than or equal to 145cm and weight less than 45kgs. History of chronic medical illness was present in 11.1% of the women and 22.2% women had faced illness during pregnancy period. (Table 2)

**Table 2:** Maternal and obstetric factors.

| Variables | Case (57)<br>n (%) | Control (114)<br>n (%) | Total (171)<br>n (%) |
| --- | --- | --- | --- |
| **Parity** |  |  |  |
| Primiparous | 38 (66.7%) | 52 (45.6%) | 90 (52.6%) |
| Multiparous | 19 (33.3%) | 62 (54.4%) | 81 (47.4%) |
| **Inter-pregnancy interval** |  |  |  |
| < 2 yrs | 1 (5.3%) | 9 (14.1%) | 10 (12.0%) |
| $\geq 2$ yrs | 18 (94.7%) | 55 (85.9%) | 73 (88.0%) |
| **Frequency of ANC visit** |  |  |  |
| < 4 visits | 5 (8.8%) | 2 (1.8%) | 7 (4.1%) |
| $\geq 4$ visits | 52 (91.2%) | 112 (98.2%) | 164 (95.9%) |
| **Hemoglobin level** |  |  |  |
| < 11 mg/dl | 14 (24.6%) | 8 (7.0%) | 22 (12.9%) |
| $\geq 11$ mg/dl | 43 (75.4%) | 106 (93.0%) | 149 (87.1%) |
| **Period of gestation** |  |  |  |
| < 37 weeks | 18 (31.6%) | 7 (6.1%) | 25 (14.6%) |
| $\geq 37$ weeks | 39 (68.4%) | 107 (93.9%) | 146 (85.4%) |
| **History of LBW** |  |  |  |
| Yes | 8 (47.1%) | 13 (20.6%) | 21 (26.3%) |
| No | 9 (52.9%) | 50 (79.4%) | 59 (73.8%) |
| **Height** |  |  |  |
| ≤ 145 cm | 7 (12.3%) | 12 (10.5%) | 19 (11.1%) |
| >145 cm | 50 (87.7%) | 102 (89.5%) | 152 (88.9%) |
| **Weight** |  |  |  |
| < 45 kg | 13 (22.8%) | 6 (5.3%) | 19 (11.1%) |
| ≥ 45 kg | 44 (77.2%) | 108 (94.7%) | 152 (88.9%) |
| **MUAC** |  |  |  |
| < 23 cm | 15 (26.3%) | 5 (4.4%) | 20 (11.7%) |
| ≥ 23 cm | 42 (73.7%) | 109 (95.6%) | 151 (88.3%) |
| **History of chronic medical illness** |  |  |  |
| Yes | 12 (21.1%) | 7 (6.1%) | 19 (11.1%) |
| No | 45 (78.9%) | 107 (93.9%) | 152 (88.9%) |
| **Illness during pregnancy** |  |  |  |
| Yes | 17 (29.8%) | 21 (18.4%) | 38 (22.2%) |
| No | 40 (70.2%) | 93 (81.6%) | 133 (77.8%) |

### Nutritional and behavioral factors of respondents

Among 171 respondents, significant portion of the respondents (34.5%) did not take any additional meal during pregnancy. Majority of the respondents were non-vegetarian (88.9%) and had taken Iron Folic Acid (IFA) for more than or equal to three months during pregnancy. Only 3.5% and 2.9% of the mothers used to smoke and consume alcohol respectively during pregnancy. Regarding duration of sleep at night 30.4% of the participants responded that they slept for less than 8 hours during pregnancy. (Table 3)

**Table 3:** Nutritional and behavioral factors.

| Variables | Case (57)<br>n (%) | Control (114)<br>n (%) | Total (171)<br>n (%) |
| --- | --- | --- | --- |
| **Additional meal during pregnancy** |  |  |  |
| No | 24 (42.1%) | 35 (30.7%) | 59 (34.5%) |
| Yes | 33 (57.9%) | 79 (69.3%) | 112 (65.5%) |
| **Type of diet** |  |  |  |
| Vegetarian | 10 (17.5%) | 9 (7.9%) | 19 (11.1%) |
| Non-vegetarian | 47 (82.5%) | 105 (92.1%) | 152 (88.9%) |
| **IFA during pregnancy** |  |  |  |
| < 3 months | 6 (10.5%) | 8 (7.0%) | 14 (8.2%) |
| ≥ 3 months | 51 (89.5%) | 106 (93.0%) | 157 (91.8%) |
| **Dietary diversity** |  |  |  |
| Inadequate (<5) | 12 (21.1%) | 5 (4.4%) | 17 (9.9%) |
| Adequate (≥5) | 45 (78.9%) | 109 (95.6%) | 154 (90.1%) |
| **Smoking habit** |  |  |  |
| Yes | 5 (8.8%) | 1 (0.9%) | 6 (3.5%) |
| No | 52 (91.2%) | 113 (99.1%) | 165 (96.5%) |
| **Alcohol consumption** |  |  |  |
| Yes | 4 (7.0%) | 1 (0.9%) | 5 (2.9%) |
| No | 53 (93.0%) | 113 (99.1%) | 166 (97.1%) |
| **Duration of sleep at night** |  |  |  |
| < 8 hours | 23 (40.4%) | 29 (25.4%) | 52 (30.4%) |
| ≥ 8 hours | 34 (59.6%) | 85 (74.6%) | 119 (69.6%) |

### Determinants of low birth weight

Bivariate and multivariate logistic regression analyses were performed to determine the association between various risk factors and birth of LBW babies. The bivariate analyses revealed that the educational status of mother, per capita income, parity, frequency of ANC visits, hemoglobin level, period of gestation, history of LBW, weight of mother before pregnancy, MUAC, history of chronic medical illness, smoking status, alcohol consumption and duration of sleep at night were significantly associated with LBW. Those variables which were significantly associated with LBW in bivariate analyses and those variables with p-value less than or equal to 0.2 were subjected to multivariate analysis using enter method and the variables having expected cell count less than 5 were excluded. Table 4 shows that illiterate women were six times more likely to give birth to LBW baby as compared to literate women (AOR: 6.32; 95% CI 1.90 to 21.05). Similarly, women whose per capita income was below poverty line were almost three times increased risk of giving birth to LBW babies (AOR: 2.89; 95% CI 1.02 to 8.18).

**Table 4:** Socio-demographic determinants of low birth weight.

| **Variables** | **p-value** | **COR (95% CI)** | **p-value** | **AOR (95% CI)** |
| --- | --- | --- | --- | --- |
| **Sex of child** |  |  |  |  |
| Female | 0.278<sup>a</sup> | 1.422 (0.75 – 2.69) |  |  |
| Male |  | Reference |  |  |
| **Ethnicity** |  |  |  |  |
| Others* | 0.072<sup>a</sup> | 0.55 (0.29 – 1.06) | 0.388 | 0.67 (0.27 – 1.66) |
| Brahmin/Chhetri |  | Reference |  | Reference |
| **Religion** |  |  |  |  |
| Non-hindu | 0.638<sup>a</sup> | 0.85 (0.42 – 1.70) |  |  |
| Hindu |  | Reference |  |  |
| **Educational status of mother** |  |  |  |  |
| Illiterate | **0.024<sup>a</sup>** | **2.77 (1.12 – 6.88)** | **0.003** | **6.32 (1.90 – 21.05)** |
| Literate |  | Reference |  | Reference |
| **Occupation of mother** |  |  |  |  |
| Home maker | 0.906<sup>a</sup> | 1.04 (0.52 – 2.08) |  |  |
| Employed |  | Reference |  |  |
| **Family size** |  |  |  |  |
| ≥ 5 members | 0.307<sup>a</sup> | 1.46 (0.70 – 3.04) |  |  |
| < 5 members |  | Reference |  |  |
| **Per capita income** |  |  |  |  |
| Below Poverty Line | **0.016<sup>a</sup>** | **2.39 (1.16 – 4.92)** | **0.046\*** | **2.89 (1.02 – 8.18)** |
| Above Poverty Line |  | Reference |  | Reference |

<sup>a</sup> = Pearson chi- square; <sup>b</sup> = Fischer's exact test, \*Significant at p-value < 0.05

**Table 5** represents that primiparous women were almost five times more likely to give birth to LBW babies as compared to multiparous women (AOR: 4.80; 95% CI 1.91 to 12.07). Anemic women had about 6 times higher odds of giving birth to LBW babies (AOR: 6.19; 95% CI 1.79 to 21.38). Preterm babies were eight times more likely to be low birth weight as compared to term infants (AOR: 8.16; 95% CI 2.42 to 27.49). Those women whose weight was less than 45 kgs before pregnancy had almost five times increased odds of giving birth to LBW babies (AOR: 4.86; 95% CI 1.02 to 23.29). Women having history of chronic medical illness were eight times more likely to have LBW baby (AOR: 8.22; 95% CI 2.25 to 29.99). Women who had suffered illness during pregnancy were at three times increased odds of having LBW baby as compared to those who had not suffered any illness during pregnancy (AOR: 3.33; 95% CI 1.20 to 9.28).

**Table 5:** Maternal and obstetric determinants of low birth weight.

| Variables | p-value | COR (95% CI) | p-value | AOR (95% CI) |
| --- | --- | --- | --- | --- |
| **Parity** |  |  |  |  |
| Primiparous | **0.009<sup>a</sup>** | **2.39 (1.13 – 4.63)** | **0.001\*** | **4.80 (1.91 – 12.07)** |
| Multiparous |  | Reference |  | Reference |
| **Inter-pregnancy interval** |  |  |  |  |
| < 2 yrs | 0.441<sup>b</sup> | 0.34 (0.04 – 2.87) |  |  |
| ≥ 2 yrs |  | Reference |  |  |
| **Frequency of ANC visit** |  |  |  |  |
| < 4 visits | **0.042<sup>b</sup>** | **5.39 (1.01 – 28.68)** |  |  |
| ≥ 4 visits |  | Reference |  |  |
| **Hemoglobin level** |  |  |  |  |
| < 11 mg/dl | **0.001<sup>a</sup>** | **4.31 (1.69 – 11.02)** | **0.004\*** | **6.19 (1.79 – 21.38)** |
| ≥ 11 mg/dl |  | Reference |  |  |
| **Period of gestation** |  |  |  |  |
| < 37 weeks | **<0.001<sup>a</sup>** | **7.06 (2.74 – 18.19)** | **0.001\*** | **8.16 (2.42 – 27.49)** |
| ≥ 37 weeks |  | Reference |  |  |
| **History of LBW** |  |  |  |  |
| Yes | **0.028<sup>a</sup>** | **3.42 (1.10 – 10.59)** |  |  |
| No |  | Reference |  |  |
| **Height** |  |  |  |  |
| ≤ 145 cm | 0.731<sup>a</sup> | 1.19 (0.44 – 3.21) |  |  |
| > 145 cm |  | Reference |  |  |
| **Weight before pregnancy** |  |  |  |  |
| < 45 kgs | **0.001<sup>a</sup>** | **5.32 (1.90 – 14.88)** | **0.048\*** | **4.86 (1.02 – 23.29)** |
| ≥ 45 kgs |  | Reference |  | Reference |
| **MUAC** |  |  |  |  |
| < 23 cm | **<0.001<sup>a</sup>** | **7.79 (2.66 – 22.76)** | 0.722 | 0.15 (0.15 – 4.06) |
| ≥ 23 cm |  | Reference |  | Reference |
| **History of chronic medical illness** |  |  |  |  |
| Yes | **0.003<sup>a</sup>** | **4.08 (1.51 – 11.03)** | **0.001\*** | **8.22 (2.25 – 29.99)** |
| No |  | Reference |  | Reference |
| **Illness during pregnancy** |  |  |  |  |
| Yes | 0.091<sup>a</sup> | 1.88 (0.89 – 3.94) | **0.021\*** | **3.33 (1.20 – 9.28)** |
| No |  | Reference |  | Reference |

<sup>a</sup> = Pearson chi- square; <sup>b</sup> = Fischer's exact test, \*Significant at p-value < 0.05 (AOR: 4.84; 95% CI 1.14 to 20.64).

**Table 6** shows that vegetarian mothers were almost five times more likely to give birth to LBW babies as compared to non-vegetarian mothers (AOR: 4.84; 95% CI 1.14 to 20.64).

**Table 6:**
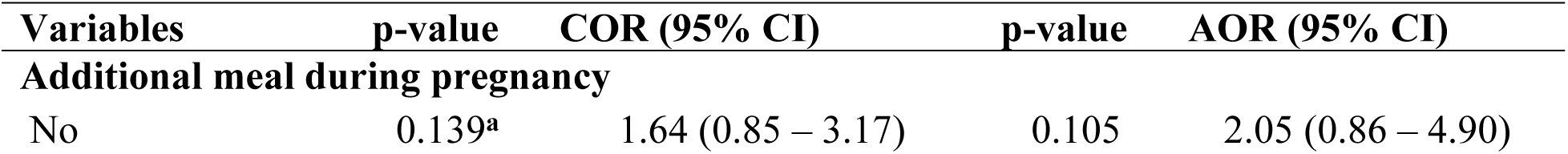
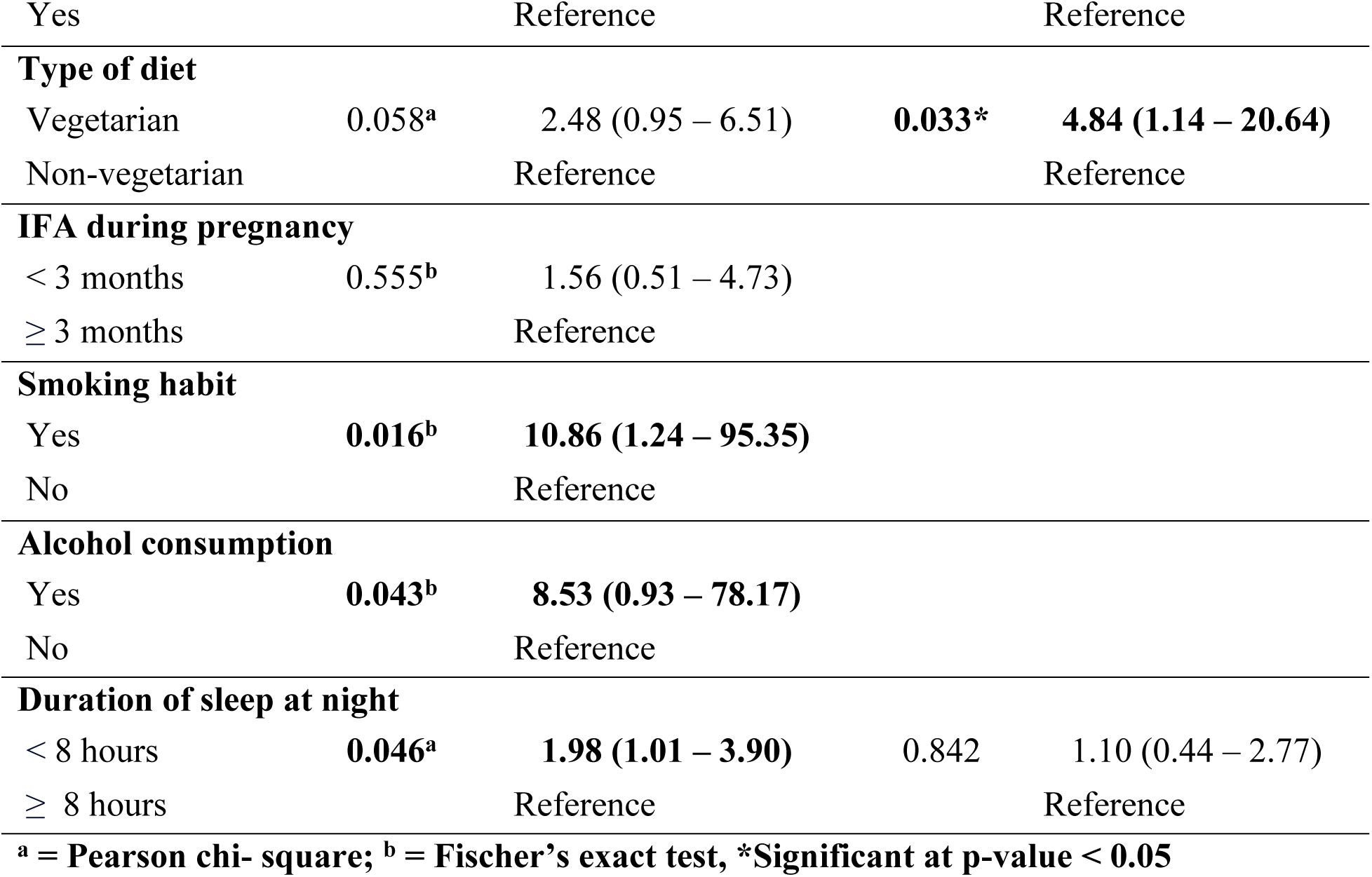
Nutritional and behavioral determinants of low birth weight.

| Variables | p-value | COR (95% CI) | p-value | AOR (95% CI) |
| --- | --- | --- | --- | --- |
| **Additional meal during pregnancy** |  |  |  |  |
| No | 0.139<sup>a</sup> | 1.64 (0.85 – 3.17) | 0.105 | 2.05 (0.86 – 4.90) |
| Yes |  | Reference |  | Reference |
| **Type of diet** |  |  |  |  |
| Vegetarian | 0.058<sup>a</sup> | 2.48 (0.95 – 6.51) | **0.033\*** | **4.84 (1.14 – 20.64)** |
| Non-vegetarian |  | Reference |  | Reference |
| **IFA during pregnancy** |  |  |  |  |
| < 3 months | 0.555<sup>b</sup> | 1.56 (0.51 – 4.73) |  |  |
| ≥ 3 months |  | Reference |  |  |
| **Smoking habit** |  |  |  |  |
| Yes | **0.016<sup>b</sup>** | **10.86 (1.24 – 95.35)** |  |  |
| No |  | Reference |  |  |
| **Alcohol consumption** |  |  |  |  |
| Yes | **0.043<sup>b</sup>** | **8.53 (0.93 – 78.17)** |  |  |
| No |  | Reference |  |  |
| **Duration of sleep at night** |  |  |  |  |
| < 8 hours | **0.046<sup>a</sup>** | **1.98 (1.01 – 3.90)** | 0.842 | 1.10 (0.44 – 2.77) |
| ≥ 8 hours |  | Reference |  | Reference |
| <sup>a</sup> = Pearson chi- square; <sup>b</sup> = Fischer's exact test, *Significant at p-value < 0.05 |  |  |  |  |

## Discussion

This study analyzed the socio-demographic factors, maternal and obstetric factors and behavioral factors of mothers with delivery of low birth weight baby. In response to educational status of mother, our study found that educational status was significantly associated with low birth weight in which illiterate mother were more prone to give birth to LBW baby as compared to literate mothers. This finding is in line with the studies conducted in Nepal, Ethiopia and Ghana which showed that illiterate mothers or mothers having lower level of education were more prone to deliver LBW baby as compared to literate mothers.^(16,20,21)^ Therefore, it would be beneficial to improve the maternal educational status in order to prevent the delivery of low birth weight baby. However, no statistical significant association was observed between mother’s educational status and LBW baby in the study conducted in Bangladesh.^(22)^ The study population in Bangladesh was relatively homogenous in terms of education, so the variation in birth outcomes between educated and uneducated mothers could be less pronounced. This homogeneity may have reduced the ability to detect significant differences in LBW rates based on education.

Our study observed a significant association between per capita income of family with LBW where the mothers whose family is below the poverty line tend to give birth to more LBW baby as compared to the mothers whose family is above poverty line. Similar findings was observed in a study conducted by Bhaskar et al. that revealed significant association between per-capita income and LBW.^(23)^ Economic status is found to be relatively well established factor for LBW baby as it is linked with overall care for the mothers during pregnancy which directly impacts the birth outcome. The present study identified parity as a significant determinant of low birth weight (LBW), with primiparous women being at a five-fold increased risk of giving birth to LBW babies compared to multiparous women. These findings are supported by studies conducted in a tertiary teaching hospital in Kathmandu, Nepal and Tamale Metropolis.^(12,24)^ Similarly, a systematic review of maternal risk factors for LBW, including a 10-year study from Korea, found that LBW rates were higher among primiparous women, which aligns with our results.^(25)^ The first-time mothers (primiparous women) might be at higher risk of giving birth to LBW baby due to factors like lack of experience in prenatal care, poorer nutritional status, or anxiety during pregnancy. However, these findings contradict those from studies conducted in Tribhuvan University Teaching Hospital (TUTH) of Nepal and selected hospitals of Indore, India, where no significant association was observed.^(26,27)^

In this study, indicating that mothers with lower hemoglobin levels <11 mg/dl had more than six times the risk of giving birth to low birth weight (LBW) babies compared to those with hemoglobin levels ≥11 mg/dl. These findings are consistent with a study conducted in Ethiopia, which found that established a link between maternal anemia and LBW.^(28)^ Other studies, including those by Kayastha et al., Sah et al., Bater et al. and Arabzadeh et al. also reported a significant association between low maternal hemoglobin levels and the incidence of LBW babies..^(12,29–31)^ However, there are studies, such as those conducted by Shrestha et al. in Lumbini Provincial Hospital and Bhaskar et al. in Eastern Nepal, that contradict these findings, as they observed no significant association between maternal hemoglobin levels and LBW births.^(16,23)^ Reduced hemoglobin levels hinder the mother’s capacity to deliver adequate oxygen and nutrients to the fetus, while also affecting her own oxygen levels and the formation of the placenta. This leads to a sustained lack of oxygen (fetal hypoxia) and inadequate nutrient supply to the fetus, which impedes healthy weight gain. Ultimately, these factors contribute to poor birth outcomes, such as low birth weight (LBW).^(32)^

Preterm birth is widely recognized as a significant risk factor for low birth weight (LBW), as babies born before 37 weeks of gestation are often not fully developed, leading to lower birth weights. In our study, women who delivered preterm were found to be 8 times more likely to have an LBW baby compared to those who carried their pregnancies to full term. The findings from our study are comparable to those reported in several other studies. For instance, research by Bansal et al., Shrestha et al., Chhetri et al. and Sutan et al. also identified preterm delivery as a significant predictor of LBW.^(8,16,33,34)^ Thus, the consistent findings across multiple studies highlight the importance of preventing preterm births to reduce the incidence of LBW and improve overall neonatal health outcomes.

Our study found that being underweight, with a pre-pregnancy weight below 45 kg, is significantly associated with higher odds of giving birth to a low birth weight baby. Similar findings were reported by Abubakari et al. in Tamale Metropolis, Sutan et al. in Malaysia, Dutta et al. at Assam Medical College in India, Devaguru et al. in Telangana, India, Andemariam et al. in Asmara and Tadese et al. in Ethiopia, all of whom identified a significant association between LBW and maternal pre-pregnancy weight.^(24,34–37)^ In contrast, studies by Mulu et al. and Wanode et al. found no significant association between maternal pre-pregnancy weight and the likelihood of delivering a LBW baby.^(20,27)^ This divergence in findings highlights the complexity of maternal health and its influence on neonatal outcomes, underscoring the need for further investigation into the role of pre-pregnancy weight in different demographic and geographic settings.

Presence of chronic medical illness among mother has an increased odds of giving birth to LBW baby in our study. This finding aligns with results from several studies, such as those by Pal et al. in West Bengal, K.C. et al. in Dang, Gebremedhin et al. in Northern Ethiopia, Mulu et al. in Addis Ababa, Ethiopia and Sutan et al. in Malaysia, which identified co-morbid conditions like hypertension as significant risk factors for LBW neonates.^(20,34,38–40)^ The biological reasoning behind this association likely stems from how hypertension affects blood flow and nutrient transfer between the mother and the fetus. In hypertensive pregnancies, reduced uteroplacental blood flow can lead to fetal growth restriction, thereby increasing the risk of LBW.^(41)^ Presence of medical illness during pregnancy was found to be significantly associated with LBW babies in our study which is in accordance with other studies conducted by Pal et al, Bhaskar et al., Toru et al. and Andemariam et al. These studies consistently found that mothers who experienced medical illnesses during pregnancy had a higher risk of delivering LBW infants compared to those who did not face any medical complications during their pregnancy.^(23,38,42,43)^ Maternal illnesses can result in poor pregnancy outcomes by affecting the mother’s overall health, nutritional status, and the ability to carry the pregnancy to full term which increases the likelihood of preterm birth, another significant risk factor for LBW.

The dietary habit of a mother during pregnancy has a significant role in the maintenance of the proper weight of newborn babies. The present study identified that vegetarian mothers had a higher risk of delivering low birth weight (LBW) babies compared to non-vegetarian mothers. This finding is in line with the study from hospital based study in Nepal conducted by Sah et al. and Canada conducted by Zulyniak MA et al. both of which reported that a plant-based diet was associated with lower birth weight.^(29,44)^ Similarly, another study by Koirala et al. in Nepal also found an increased risk of LBW among vegetarian mothers, though the association was not statistically significant.^(45)^ Animal-based foods, which are high in iron and essential hemopoietic nutrients such as vitamin B12 and folic acid, contribute to raising hemoglobin levels in pregnant women.^(46)^ Adequate intake of these nutrients reduces the likelihood of low birth weight in newborns by supporting proper fetal growth.

Smoking and alcohol consumption during pregnancy are an established significant risk factors for low birth weight (LBW) babies, adversely affecting both maternal and fetal health. This present study revealed that both smoking and alcohol consumption during pregnancy posed a greater risk of giving birth to LBW babies. These findings are supported by systematic review and meta-analysis of 55 cohort studies from 1986 to 2020 conducted by Kun Di et al. and an umbrella review conducted by Arabzadeh et al.^(31,47)^ Other studies such as those conducted by Kayastha et al. in Nepal, Taywade et al. in Wardha district of India and Devaguru et al. in Telangana, India also found the results that are consistent to the results of our study.^(12,36,48)^ Despite being an established risk factors for pregnancy, our study may not hold the statistical power to predict the outcomes since the number of smokers and alcohol consumers were very less in our study.

This study has several limitations. As the study is of retrospective nature, participants may fail to remember the factors that have happened in the past. Thus, it may result in recall bias. The study was conducted in a single government hospital within the Kathmandu Valley so the findings primarily reflect mothers delivering in similar institutional settings and may not capture all community-level variations. Since the data were collected using face-to-face interviews, it might have resulted in social desirability bias in some self-reported behaviors like dietary practices, smoking and alcohol consumption. The relatively small number of participants who reported smoking or alcohol consumption also limited the statistical power to detect strong associations with these factors. Despite these limitations, the study provides robust evidence on modifiable maternal and socio-demographic determinants of low birth weight, which can inform both hospital-based and community-level interventions in Nepal.

## Conclusions

The research identified key maternal risk factors independently linked to LBW, including the per capita income below poverty line, higher parity, anemia (low hemoglobin levels), preterm delivery (gestational age less than 37 weeks), being underweight before pregnancy, history of chronic medical conditions, illnesses during pregnancy and being vegetarian. Despite an initial analysis pointing to additional factors such as mid-upper arm circumference (MUAC), and diverse diet during pregnancy, these were not found to be significant when adjusted for confounding variables. The findings underscore the critical role of maternal health, nutrition, and prenatal care in preventing LBW. Interventions that address maternal education, nutrition, regular antenatal visits, and timely management of maternal illnesses could play a pivotal role in reducing the incidence of LBW, ultimately improving neonatal outcomes.

## Data Availability

The datasets generated and/or analysed during the current study are available in the figshare repository, https://doi.org/10.6084/m9.figshare.29815940.v1

## Additional information

## Ethics approval and consent to participate

Ethical approval was obtained from Institutional Review Committee (IRC), BP Koirala Institute of Health Sciences, Dharan (Reference number: IRC/30/081/82). Permission to conduct the study was also obtained from Paropakar Maternity and Women’s Hospital, Kathmandu. Written informed consent was obtained from all participants prior to data collection. Confidentiality and anonymity of the respondents were maintained throughout the study by using unique participant codes instead of personal identifiers.

## Author contribution

**Conceptualization:** Sadiksha Pokhrel, Paras Kumar Pokharel, Anup Ghimire, Birendra Kumar Yadav

**Data curation:** Sadiksha Pokhrel, Birendra Kumar Yadav, Swastika Poudel, Akshaya Acharya

**Formal analysis:** Sadiksha Pokhrel, Swastika Poudel, Birendra Kumar Yadav

**Methodology:** Sadiksha Pokhrel, Anup Ghimire, Birendra Kumar Yadav, Avaniendra Chakravartty, Swastika Poudel

**Project Administration:** Paras Kumar Pokharel, Sadiksha Pokhrel, Anup Ghimire

**Resources:** Laxmiswori Prajapati, Sadiksha Pokhrel

**Supervision:** Paras Kumar Pokharel, Anup Ghimire, Laxmiswori Prajapati

**Validation:** Sadiksha Pokhrel, Paras Kumar Pokharel, Anup Chimire, Birendra Kumar Yadav, Avaniendra Chakravartty, Laxmiswori Prajapati

**Visualization:** Sadiksha Pokhrel

**Writing original draft:** Sadiksha Pokhrel, Swastika Poudel

**Writing-review and editing:** Sadiksha Pokhrel, Swastika Poudel, Akshaya Acharya, Paras Kumar Pokharel

## Acknowledgements

We would like to express our sincere gratitude to the BP Koirala Institute of Health Sciences (BPKIHS) for academic guidance and support throughout the course of this study. Special thanks are extended to the IRC-BPKIHS and Paropakar Maternity and Women’s Hospital, Kathmandu, for granting permission to conduct the research. We are also grateful to all the participants who kindly consented to be part of this study.

## References

1. World Health Organization. International Statistical Classification of Diseases and Related Health Problems - 10th revision. WHO [Internet]. 2016;1. Available from: https://apps.who.int/iris/bitstream/handle/10665/246208/9789241549165-V1-eng.pdf

2. Pal A, Manna S, Das B, Dhara PC. The risk of low birth weight and associated factors in West Bengal, India: a community based cross-sectional study. Egypt Pediatr Assoc Gaz. 2020 Sep 1;68(1):27.

3. Jana A, Saha UR, Reshmi RS, Muhammad T. Relationship between low birth weight and infant mortality: evidence from National Family Health Survey 2019-21, India. Arch Public Health. 2023 Feb 21;81(1):28.

4. Low Birth Weight - Health Encyclopedia - University of Rochester Medical Center [Internet]. [cited 2023 Dec 28]. Available from: https://www.urmc.rochester.edu/encyclopedia/content.aspx?contenttypeid=90&contentid=p02382

5. default - Stanford Medicine Children’s Health [Internet]. [cited 2023 Dec 28]. Available from: https://www.stanfordchildrens.org/en/topic/default?id=low-birthweight-90-P02382

6. Cleveland Clinic [Internet]. [cited 2023 Dec 28]. Low Birth Weight. Available from: https://my.clevelandclinic.org/health/diseases/24980-low-birth-weight

7. UNICEF DATA [Internet]. [cited 2023 Dec 28]. Low birthweight. Available from: https://data.unicef.org/topic/nutrition/low-birthweight/

8. Bansal P, Garg S, Upadhyay HP. Prevalence of low birth weight babies and its association with socio-cultural and maternal risk factors among the institutional deliveries in Bharatpur, Nepal. Asian J Med Sci. 2018 Dec 9;10(1):77–85.

9. Marete I, Ekhaguere O, Bann CM, Bucher SL, Nyongesa P, Patel AB, et al. Regional trends in birth weight in low- and middle-income countries 2013–2018. Reprod Health. 2020 Dec;17(S3):176.

10. Feresu SA, Harlow SD, Woelk GB. Risk Factors for Low Birthweight in Zimbabwean Women: A Secondary Data Analysis. Thorne C, editor. PLOS ONE. 2015 Jun 26;10(6):e0129705.

11. Prajapati R, Shrestha S, Bhandari N. Prevalence and Associated Factors of Low Birth Weight among Newborns in a Tertiary Level Hospital in Nepal. Kathmandu Univ Med J. 2018;16(1)(61):49–52.

12. Kayastha P, Manandhar SR. Incidence and Risk Factors of Low Birth Weight Among Babies Delivered at Tertiary Level Teaching Hospital in Nepal. Med J Shree Birendra Hosp. 2019 Jul 12;18(2):29–35.

13. Global Nutrition Report | Country Nutrition Profiles - Global Nutrition Report [Internet]. [cited 2023 Dec 29]. Available from: https://globalnutritionreport.org/resources/nutrition-profiles/asia/south-eastern-asia/

14. Nepal Demographic and Health Survey 2022 [Internet]. Ministry of Health and Population; 2022. Available from: https://dhsprogram.com/pubs/pdf/FR379/FR379.pdf

15. Annual Report 2077/78 (2020/21) [Internet]. Department of Health Services; 2020. Available from: http://dohs.gov.np/annual-report-fy-2077-78-2019-20/

16. Shrestha S, Shrestha S, Shakya Shrestha U, Gyawali K. Predictors of Low Birth Weight at Lumbini Provincial Hospital, Nepal: A Hospital-Based Unmatched Case Control Study. Adv Prev Med. 2020 Mar 26;2020:1–7.

17. Office of the Prime Minister and Council of Ministers. Nepal Living Standard Survey IV 2022-23 [Internet]. National Statistics Office; 2024. Available from: https://nepalindata.com/resource/NEPAL-LIVING-STANDARDS-SURVEY-IV-2022-23/

18. Guide to Anthropometry: A Practical Tool for Program Planners, Managers, and Implementers | Food and Nutrition Technical Assistance III Project (FANTA) [Internet]. 2018 [cited 2024 Jan 3]. Available from: https://www.fantaproject.org/tools/anthropometry-guide

19. FAO. Minimum dietary diversity for women: An updated guide to measurement - from collection to action [Internet]. Rome, Italy: FAO; 2021 [cited 2024 Jan 3]. 176 p. Available from: https://www.fao.org/documents/card/en?details=cb3434en

20. Baye Mulu G, Gebremichael B, Wondwossen Desta K, Adimasu Kebede M, Asmare Aynalem Y, Bimirew Getahun M. Determinants of Low Birth Weight Among Newborns Delivered in Public Hospitals in Addis Ababa, Ethiopia: Case-Control Study. Pediatr Health Med Ther. 2020 Mar;Volume 11:119–26.

21. Adam Z, Ameme DK, Nortey P, Afari EA, Kenu E. Determinants of low birth weight in neonates born in three hospitals in Brong Ahafo region, Ghana, 2016-an unmatched case-control study. BMC Pregnancy Childbirth. 2019 Dec;19(1):174.

22. Siddiqi MdNA, Muyeed A, Haque MdN, Abdul Goni Md, Shadhana SC. Low Birth Weight of Newborns and Its Association with Demographic and Socio-economic Determinants: Findings from Multiple Indicator Cluster Survey (MICS) Bangladesh 2019. Int J Health Stud [Internet]. 2021 Feb 17 [cited 2024 Jan 1];7(1). Available from: 10.22100/ijhs.v7i1.837

23. Bhaskar RK, Deo KK, Neupane U, Chaudhary Bhaskar S, Yadav BK, Pokharel HP, et al. A Case Control Study on Risk Factors Associated with Low Birth Weight Babies in Eastern Nepal. Int J Pediatr. 2015;2015:1–7.

24. Abubakari A, Asumah MN, Abdulai NZ. Effect of maternal dietary habits and gestational weight gain on birth weight: an analytical cross-sectional study among pregnant women in the Tamale Metropolis. Pan Afr Med J [Internet]. 2023 [cited 2024 Sep 24];44. Available from: https://www.panafrican-med-journal.com/content/article/44/19/full

25. Siti Lutfia, Sofia Al Farizi. Maternal risk factors for low birth weight infants: A systematic review study. World J Adv Res Rev. 2024 Jan 30;21(1):1548–54.

26. Sharma SR, Giri S, Timalsina U, Bhandari SS, Basyal B, Wagle K, et al. Low Birth Weight at Term and Its Determinants in a Tertiary Hospital of Nepal: A Case-Control Study. Baud O, editor. PLOS ONE. 2015 Apr 8;10(4):e0123962.

27. Deepika Wanode, Amar Yadav. Assess the maternal factors associated with Birth weight of newborn. World J Adv Res Rev. 2023 Nov 30;20(2):797–803.

28. Ejeta Chibsa S, Adem Hussen M, Bayisa K, Tefera Kefeni B. Determinants of low birth weight among newborns delivered at Mettu Karl comprehensive specialized hospital, southwest Ethiopia: a case–control study. Sci Rep. 2024 Feb 22;14(1):4399.

29. Sah SK, Sunuwar DR, Baral JR, Singh DR, Chaudhary NK, Gurung G. Maternal hemoglobin and risk of low birth weight: A hospital-based cross-sectional study in Nepal. Heliyon. 2022 Dec;8(12):e12174.

30. Bater J, Lauer JM, Ghosh S, Webb P, Agaba E, Bashaasha B, et al. Predictors of low birth weight and preterm birth in rural Uganda: Findings from a birth cohort study. Wilunda C, editor. PLOS ONE. 2020 Jul 13;15(7):e0235626.

31. Arabzadeh H, Doosti-Irani A, Kamkari S, Farhadian M, Elyasi E, Mohammadi Y. The maternal factors associated with infant low birth weight: an umbrella review. BMC Pregnancy Childbirth. 2024 Apr 25;24(1):316.

32. Ali SA, Tikmani SS, Saleem S, Patel AB, Hibberd PL, Goudar SS, et al. Hemoglobin concentrations and adverse birth outcomes in South Asian pregnant women: findings from a prospective Maternal and Neonatal Health Registry. Reprod Health. 2020 Nov;17(S2):154.

33. Chhetri M, Tripathi G, Joshi R, Koirala S, Chapagain S, Subedi M. BIRTH WEIGHT AND ITS ASSOCIATED FACTORS AMONG LIVE BIRTHS AT CHITWAN MEDICAL COLLEGE, NEPAL. J Chitwan Med Coll. 2022 Jan 19;11(4):28–31.

34. Sutan R, Mohtar M, Mahat AN, Tamil AM. Determinant of Low Birth Weight Infants: A Matched Case Control Study. Open J Prev Med. 2014;04(03):91–9.

35. Dutta A, Sonowal BB, Dhungel L. Maternal profile of mothers giving birth to low birth weight babies: A hospital based cross-sectional study. New Indian J OBGYN. 2024 Apr;10(2):314–9.

36. Devaguru A, Gada S, Potpalle D, Dinesh Eshwar M, Purwar D. The Prevalence of Low Birth Weight Among Newborn Babies and Its Associated Maternal Risk Factors: A Hospital-Based Cross-Sectional Study. Cureus [Internet]. 2023 May 5 [cited 2024 Sep 26]; Available from: https://www.cureus.com/articles/143738-the-prevalence-of-low-birth-weight-among-newborn-babies-and-its-associated-maternal-risk-factors-a-hospital-based-cross-sectional-study

37. Tadese M, Minhaji AS, Mengist CT, Kasahun F, Mulu GB. Determinants of low birth weight among newborns delivered at Tirunesh Beijing General Hospital, Addis Ababa, Ethiopia: a case-control study. BMC Pregnancy Childbirth. 2021 Dec;21(1):794.

38. Pal A, Manna S, Das B, Dhara PC. The risk of low birth weight and associated factors in West Bengal, India: a community based cross-sectional study. Egypt Pediatr Assoc Gaz. 2020 Dec;68(1):27.

39. K. C. A, Basel PL, Singh S. Low birth weight and its associated risk factors: Health facility-based case-control study. Pradhan PMS, editor. PLOS ONE. 2020 Jun 22;15(6):e0234907.

40. Gebremedhin M, Ambaw F, Admassu E, Berhane H. Maternal associated factors of low birth weight: a hospital based cross-sectional mixed study in Tigray, Northern Ethiopia. BMC Pregnancy Childbirth. 2015 Dec;15(1):222.

41. Luger RK, Kight BP. Hypertension In Pregnancy. In: StatPearls [Internet]. Treasure Island (FL): StatPearls Publishing; 2025 [cited 2025 Jul 23]. Available from: http://www.ncbi.nlm.nih.gov/books/NBK430839/

42. Toru T, Anmut W. Assessment of Low Birth Weight and Associated Factors Among Neonates in Butajira General Hospital, South Ethiopia, Cross Sectional Study, 2019. Int J Pediatr. 2020 Aug 1;2020:1–6.

43. Andemariam Z, Karuppusamy S, Ghebreyohannes G, Teages E, Andemichael G. Determinants for low birth weight in Asmara, Eritrea: A maternity hospital-based study. Public Health Toxicol. 2023 Dec 28;3(4):1–9.

44. Zulyniak MA, De Souza RJ, Shaikh M, Desai D, Lefebvre DL, Gupta M, et al. Does the impact of a plant-based diet during pregnancy on birth weight differ by ethnicity? A dietary pattern analysis from a prospective Canadian birth cohort alliance. BMJ Open. 2017 Nov;7(11):e017753.

45. Koirala AK, Bhatta DN. Low-birth-weight babies among hospital deliveries in Nepal: a hospital-based study. Int J Womens Health. 2015 Jun;581.

46. Sunuwar DR, Sangroula RK, Shakya NS, Yadav R, Chaudhary NK, Pradhan PMS. Effect of nutrition education on hemoglobin level in pregnant women: A quasi-experimental study. Glover-Amengor M, editor. PLOS ONE. 2019 Mar 21;14(3):e0213982.

47. Di HK, Gan Y, Lu K, Wang C, Zhu Y, Meng X, et al. Maternal smoking status during pregnancy and low birth weight in offspring: systematic review and meta-analysis of 55 cohort studies published from 1986 to 2020. World J Pediatr. 2022 Mar 1;18(3):176–85.

48. Taywade ML, Pisudde PM. Study of sociodemographic determinants of low birth weight in Wardha district, India. Clin Epidemiol Glob Health. 2017 Mar;5(1):14–20.

